# Altered increase in STAT1 expression and phosphorylation in severe COVID-19

**DOI:** 10.1101/2021.08.13.21262006

**Authors:** Hector Rincon-Arevalo, Arman Aue, Jacob Ritter, Franziska Szelinski, Dmytro Khadzhynov, Daniel Zickler, Luisa Stefanski, Andreia C. Lino, Sixten Körper, Kai-Uwe Eckardt, Hubert Schrezenmeier, Thomas Dörner, Eva V. Schrezenmeier

## Abstract

The interferon pathway represents a key antiviral defense mechanism and is being considered as a therapeutic target in COVID-19. Both, substitution of interferon and blocking interferon signaling through JAK STAT inhibition to limit cytokine storms have been proposed. However, little is known so far about possible abnormalities in STAT signaling in immune cells during SARS-CoV-2 infection. In the current study, we investigated downstream targets of interferon signaling, including STAT1, pSTAT1 and 2 and IRF1, 7 and 9 by flow cytometry in 30 patients with COVID-19, 17 with mild and 13 with severe infection. We report an upregulation of STAT1 and IRF9 in mild and severe COVID-19 cases, which correlated with the IFN-signature assessed by Siglec-1 (CD169) expression on peripheral monocytes. Most interestingly, Siglec-1 and STAT1 in CD14+ monocytes and plasmablasts showed lower expression among severe COVID-19 cases compared to mild cases. Contrary to the baseline whole protein STAT1 expression, the phosphorylation of STAT1 was enhanced in severe COVID-19 cases, indicating a dysbalanced JAK STAT signaling that fails to induce transcription of interferon stimulated response elements (ISRE). This abnormality persisted after IFN-α and IFN-γ stimulation of PBMCs from patients with severe COVID-19. The data suggest impaired STAT1 transcriptional upregulation among severely infected patients which may represent a potential predictive biomarker and may allow stratification of patients for certain interferon-pathway targeted treatments.

## 1. Introduction

Following viral infection, a complex regulatory system of innate and adaptive immune mechanisms is initiated to defend against viral invasion. One of many responses to viral infection is the induction of the pleiotropic cytokine interferon (IFN) [1]. It acts as a key link between the innate and adaptive immune system. IFN-α (type I IFN) is mainly secreted by plasmacytoid dendritic cells (pDCs), while IFN-γ (type II IFN) is predominantly produced by natural killer cells and by certain T cells and macrophages. Both, type I and type II IFN, have diverse but complementary antiviral effects such as induction of apoptosis and activation of macrophages, natural killer (NK) cells as well as B and T lymphocytes [2, 3].

Regarding severe acute respiratory syndrome coronavirus type 2 (SARS-CoV-2) infection, IFN related antiviral response attracted attention since inborn errors of type I IFN and the presence of autoantibodies against type I IFN were found to be associated with a severe course of the disease [4, 5]. Roughly 10% of coronavirus disease 2019 (COVID-19) patients with severe pneumonia produce neutralizing autoantibodies against IFNα, IFNω, or both, while patients with no or mild disease have no detectable autoantibodies [4]. Further, it has been shown that patients with severe COVID-19 have a highly impaired IFN type I signature, with reduced IFN-α production and activity [6].

Type I IFNs signal through Janus kinase (JAK) and signal transducer and activator of transcription (STAT) pathway and thereby stimulate gene expression. After specific binding to the IFN-α receptor (IFNAR), consisting of two chains (IFNAR1 and IFNAR2), autophosphorylation of the newly formed complex leads to phosphorylation of the receptor-associated JAK1 and TYK2 [7, 8]. Subsequently, further phosphorylation of cytoplasmatic STAT1 and STAT2 induces its dimerization and interaction with IFN regulatory factor 9 (IRF9), a strong enhancer of translocation to the nucleus [9-11]. The activation of STAT1 is achieved by tyrosine phosphorylation on Y701 that is followed by nuclear accumulation [12].

Formation of a complex called interferon-stimulated gene factor 3 (ISGFR3) binding to IFN-stimulated response elements (ISRE) finally results in stimulation of gene expression [12]. Likewise the same sequence of biochemical processes leads to the transcriptional influence of IFN-γ (type II IFN), involving the IFN-γ receptor (IFNGR), JAK1 and JAK2, and dimerization of the homodimer STAT1 binding to IFN-γ-activated site (GAS) [1]. Figure 1A schematically depicts JAK/STAT signaling. Although it has been shown that the virus proteins NSP5, ORF7a, N and ORF6 are able to directly interfere with JAK STAT signaling by inhibiting translocation of STATs or inhibiting their phosphorylation in SARS-CoV-2 infected cultured cells [13, 14], it is not fully understood how JAK STAT signaling is altered in immune cells and if a dysbalance in JAK STAT signaling may contribute to disease severity.

**Figure 1.**
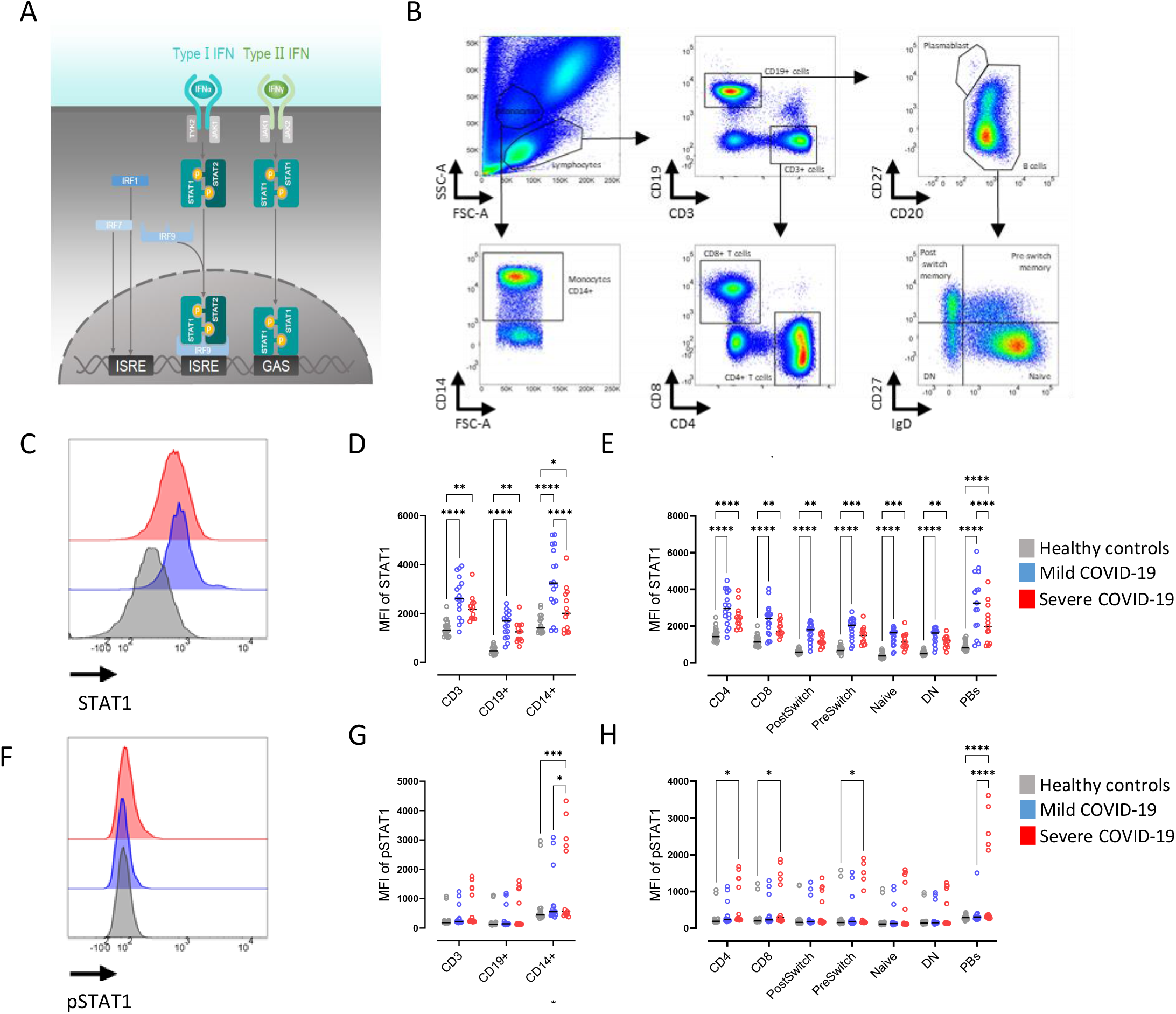
Reduced STAT1 expression in severe COVID-19 patients. (A) Schematic depiction of JAK STAT signaling. (B) Gating strategy on whole blood flow cytometry for IgD+CD27-(Naïve), IgD+CD27+ (PreSwitch), IgD-CD27+ (PostSwitched) and IgD-CD27-(Double Negative, DN) as well as CD4+ and CD8+ T cells, (C) Representative histograms of baseline expression of STAT1 on B cells from healthy controls (grey), mild (blue) and severe (red) COVID-19 patients. (D) Median fluorescence intensity (MFI) of STAT1 in CD3+, CD19+ and CD14+ cells. (E) MFI of STAT1 in T and B cell subsets (as described in B). (F) Representative histograms of baseline expression of pSTAT1 on B cells from healthy controls (grey), mild (blue) and severe (red) COVID-19 patients. (G) MFI of STAT1 in CD3+, CD19+ and CD14+ cells. (H) MFI of pSTAT1 in T and B cell subsets (as described in B). Median and data from healthy controls (n=20), mild COVID-19 (n=17) and severe COVID-19 (n=13) patients. Two way ANOVA with Sidack post-test. *p<0.05, **p<0.01, ***p<0.001 and ****p<0.0001.

Here we report an increased STAT1 expression in mild and severe COVID-19 patients compared to controls. Severe COVID cases showed lower STAT1 expression than mild patients, accompanied by elevated phosphorylation of STAT1 at the pY701 phosphosite suggesting a disturbance of signal transduction related to impaired STAT1 transcription, which is not surmountable by additional IFN stimulation.

## 2. Methods

### 2.1. Study participants

Peripheral blood samples (EDTA anti-coagulated, BD vaccutainer system, BD Diagnostics, Franklin Lakes, NJ, USA) from 20 healthy controls and 30 COVID patients were analyzed, 17 with mild (WHO 8-point ordinal scale 1 and 2) and 13 with a severe course of the disease (WHO 8-point ordinal scale ≥ 4) [15]. Donor information is summarized in Table 1. Severe cases were participants in the CAPSID trial that investigated convalescent plasma therapy (EudraCT2020-001210-38; NCT04433910I). Samples for this study were collected at baseline prior to administration of the investigation drug. All participants or their legal representatives gave written informed consent according to the approval of the ethics committee at the Charité University Hospital, Berlin (EA2/066/20 Pa-Covid-19 and University of Ulm (CAPSID trial (115/20 and 488/20) [16, 17].

**Table 1:** Patient characteristics.

|  | Mild COVID (n=17) | Severe COVID (n=13) | Healthy controls (n=20) |
| --- | --- | --- | --- |
| **Age (median, IQR)** | 47.4 (34.0; 57.0) | 61 (53.5; 69.5) | 35 (29.0;47.0) |
| **Female** | 7 | 1 | 14 |
| **Days post symptom onset** | 7 (10; 5.5) | 10 (8.5; 15) |  |
| **Concomitant antibiotic treatment** | 0 | 6 |  |
| **Immunosuppressive treatment before COVID** | 4 | 1 |  |
| **Dexamethasone** | 0 | 12 |  |
| **Tocilizumab** | 0 | 1 |  |
| **Remdesivir** | 0 | 1 |  |
| **CRP mg/dl** | n.a. | 116 (85.75; 188) |  |
| **WHO Ordinal scale 1** | 2 |  |  |
| **WHO Ordinal scale 2** | 15 |  |  |
| **WHO Ordinal scale 5** |  | 6 |  |
| **WHO Ordinal scale 6** |  | 1 |  |
| **WHO Ordinal scale 8** |  | 6 |  |

### 2.2. Analytical methods and flow cytometry

Intracellular phenotyping of STAT1, pSTAT1, pSTAT2, IRF1 and IRF7 levels in B and T cells was conducted as previously published [18]. Briefly, peripheral whole blood (500 µl) was lysed and fixed the same day of acquisition using 5 ml BD Phosflow Lyse/Fix Buffer (BD Biosciences, San Jose, CA, USA, 10 min, 37°C) considering the manufacturer’s protocol (mix of 1:5 in aqua dest). After two washing steps using ice-cold PBS Dulbecco (Biochrom GMBH, Berlin, Germany) (530 g, 8 min, 4°C), permeabilization was performed using 200 µl BD Perm Buffer II (BD Biosciences, San Jose, CA, USA; 12h, −20°C), followed by overnight storage at −20°C. Next, cells were washed twice with PBS Dulbecco containing 0.5% BSA/EDTA (530 xg, 8 min, 4°C) and resuspended in 50 µl of PBS with 20% Brilliant Buffer (BD Horizon, San Jose, CA, USA). After incubation with 2.5 µl of FcR blocking reagent (Miltenyi Biotec Bergisch Gladbach, NRW, Germany) for 5 min, staining for 1 h and subsequent flow cytometry analysis were carried out at the same day.

All flow cytometry analyses were performed using a BD FACS Fortessa (BD Biosciences, Franklin Lakes, NJ, USA). To ensure comparable mean fluorescence intensities (MFIs) over time of the analyses, Cytometer Setup and Tracking beads (CST beads, BD Biosciences, Franklin Lakes, NJ, USA) and Rainbow Calibration Particles (BD Biosciences, Franklin Lakes, NJ, USA) were used. For flow cytometric analysis, the following fluorochrome-labeled antibodies were used: BUV395 anti-CD14 (BD, clone M5E2, 1:50), PE-Cy7 anti-CD3 (BD, clone UCHT1, 1:100), BV510 anti-CD4 (BD, clone SK3, 1:50), BUV737 anti-CD8 (BD, clone SK1, 1:500), BV711 anti-CD19 (BD, clone SJ25C1,1:25), BV421 anti-CD20 (BD, clone 2H7, 1:25), BV786 anti-CD27 (BD, clone L128, 1:50), PE-CF594 anti-IgD (Biolegend, San Diego, CA, USA, clone IA6-2, 1:500) for basic immunophenotyping of cell populations of interest. Quantitative analysis was done using the following intracellular fluorochrome-labeled antibodies: PE anti-STAT1 (BD, clone 1/Stat1, 1:10), FITC anti-pSTAT1 (BD, clone 4a, 3:20), AF647 anti-pSTAT2 (R&D Systems, clone 1021D, 1:5), PE anti-IRF1 (BD, clone 20/IRF-1, 1:20), AF647 anti-IRF7 (BD, clone K47-671, 1:10). For IRF9 analysis an unconjugated IRF9 antibody (Thermo Fisher, isotype rabbit IgG, clone 14H9L22, 1:100) was applied. After staining and washing cells, a secondary antibody was used for specific binding of IRF9 (Jackson Immuno Research, isotype donkey anti rabbit IgG, polyclonal, 1:100). Number of absolute B cells was measured with Trucount (BD) and samples were processed according to the manufacturer’s instruction. As a control, at least one healthy control sample was processed simultaneously with patients’ samples. Siglec-1 (CD169) expression analysis on CD14+ monocytes was performed at baseline as previously described [19].

### 2.3. Isolation of peripheral blood mononuclear cells (PBMCs)

Peripheral blood mononuclear cells (PBMCs) were obtained by density gradient centrifugation using Ficoll-Paque PLUS (GE Healthcare Bio-Sciences, Chicago, IL, USA).

### 2.4. Functional analysis of IFNα and IFNγ signaling pathways

In order to evaluate the functional cellular responsiveness upon IFN stimuli, stimulation experiments were carried out as previously described [18]. In brief, we suspended isolated PBMCs in RPMI medium (GlutaMAX, Life Technologies, Paisley, UK) and stimulated with IFNα2a (100 ng/ml for 5 min and 5 ng/ml 48h) (Recombinant Human, Milteny, Germany) or IFNγ1b (100 ng/ml for 5 min and 5 ng/ml 48h) (Recombinant Human, Milteny, Germany). Cells were then harvested, washed, lysed, permeabilized and stained using the same staining as mentioned above followed by flow cytometry.

### 2.5. Statistical analysis

Flow cytometry data were analysed using FACSDiva software (Becton Dickinson, Franklin Lakes, NJ, USA) and FlowJo (version 10, TreeStar, Ashland, OR, USA). For graphical and statistical analysis, GraphPad Prism (version 7.00, GraphPad Software, La Jolla, CA, USA) was used. Mann–Whitney tests were used for the comparison of two groups. For multiple group comparison, two-way ANOVA with Šidák’s post-test for multiple comparison. Spearman correlation coefficient was calculated to detect possible associations between parameters or disease activity, respectively. P-values < 0.05 were considered significant. Correlation matrix was calculated using base R and corrplot package (R Foundation for Statistical Computing) using Spearman method.

## 3. Results

### 3.1. Cohort characteristics

In the current cohort, we included 17 out-patients with mild COVID-19 during their quarantine (WHO 8-point ordinal scale 1 and 2) and 13 hospitalized patients treated for severe COVID-19 pneumonia on an Intensive Care Unit (ICU) (WHO 8-point ordinal scale ≥ 4) (Table 1) [15]. Age and days post symptom onset were not significantly different between both groups, while more patients with mild COVID-19 were female. Among patients with severe disease manifestation, 6 patients subsequently died because of COVID-19.

### 3.2. Reduced STAT1 expression in patients with severe COVID-19

Since severe COVID-19 has a highly impaired IFN type I signature, with especially reduced IFN-α production and activity [20], we initially asked how the major effector downstream targets of type I IFN are affected among patients with COVID-19. We found a significantly increased STAT1 protein expression in all analyzed cell subsets from whole blood analysis, including T cells (CD4^+^ and CD8^+^), B cells (IgD^+^CD27^-^ (Naïve), IgD^+^CD27^+^ (PreSwitch), IgD^-^CD27^+^ (PostSwitched) and IgD^-^CD27^-^ (Double Negative, DN)), plasmablasts (CD20^low^CD27^high)^ and monocytes (CD14^+^) from all patients with COVID-19 compared to healthy controls (Figure 1 C-E). Most notably, reduced STAT1 was observed in severe COVID-19 cases compared to mild COVID-19 cases in particular in CD14+ monocytes and plasmablasts, respectively (Fig 1D and Fig 1E).

Since STAT1 signaling is mainly regulated through phosphorylation, we were interested in the phosphorylated form of STAT1. Baseline pSTAT1 was significantly increased in monocytes, CD4+ and CD8+ T cells, pre-switched B cells (IgD^+^CD27^+^) and plasmablasts from severe COVID-19 patients compared to healthy controls (Figure 1F-H).

To better understand the relation between the expression of full protein STAT1 and phosphorylated protein (pSTAT1), the ratio of pSTAT1/STAT1 was obtained. A reduced ratio of pSTAT1/STAT1 was observed in plasmablasts and CD14^+^ cells from COVID-19 patients with mild disease compared to those with severe disease (Figure S1A). No differences in pSTAT2 levels were observed among study groups (Figure S1B). These results suggest that mildly affected COVID-19 patients increased their STAT1 expression, but not their detectable levels of phosphorylation, while severely affected COVID-19 patients showed a greater increase of phosphorylation in relation to the increase of total STAT1 expression. Thus, there is a substantial impairment to increase STAT1 transcription in severe COVID-19 that especially affects CD14+ monocytes and plasmablasts.

### 3.3. Reduced IRF9 expression in patients with severe COVID-19

STAT1 is part of interferon mediated viral response, representing a key component of complexes like ISGF3 and GAS, responsible for amplified interferon-mediated signals [12]. To better understand the nature of STAT1 alteration in patients with COVID-19, IRF9 (components of ISGF3 and GAS complexes) was evaluated. An increased IRF9 expression was found in CD8^+^ T cells, IgD^+^CD27^-^, IgD^+^CD27^+^, IgD^-^CD27^+^ and IgD^-^CD27^-^ B cells, PB and CD14^+^ cells from patients with mild COVID-19 compared to healthy controls (Fig. 2B, C). Similar to STAT1, severe COVID-19 was characterized by reduced IRF9 expression compared to mild COVID-19, in PB and CD14+ cells (Figure 2 A-C). Patients with severe COVID-19 had lower IRF9 expression compared to mild cases, consistent with a reduced STAT1 expression. Regarding the expression of IRF1, a molecule well known to directly bind and impact the ISRE [12], we found no difference between healthy controls and patients with COVID-19 irrespective of severity in T cells, and IgD^+^CD27^-^, IgD^+^CD27^+^, IgD^-^CD27^+^ and IgD^-^CD27^-^ B cells while in CD14+ monocytes and plasmablasts, IRF1 was significantly increased among patients with mild or severe COVID-19 compared to healthy controls (Fig. 2 D-F).

**Figure 2.**
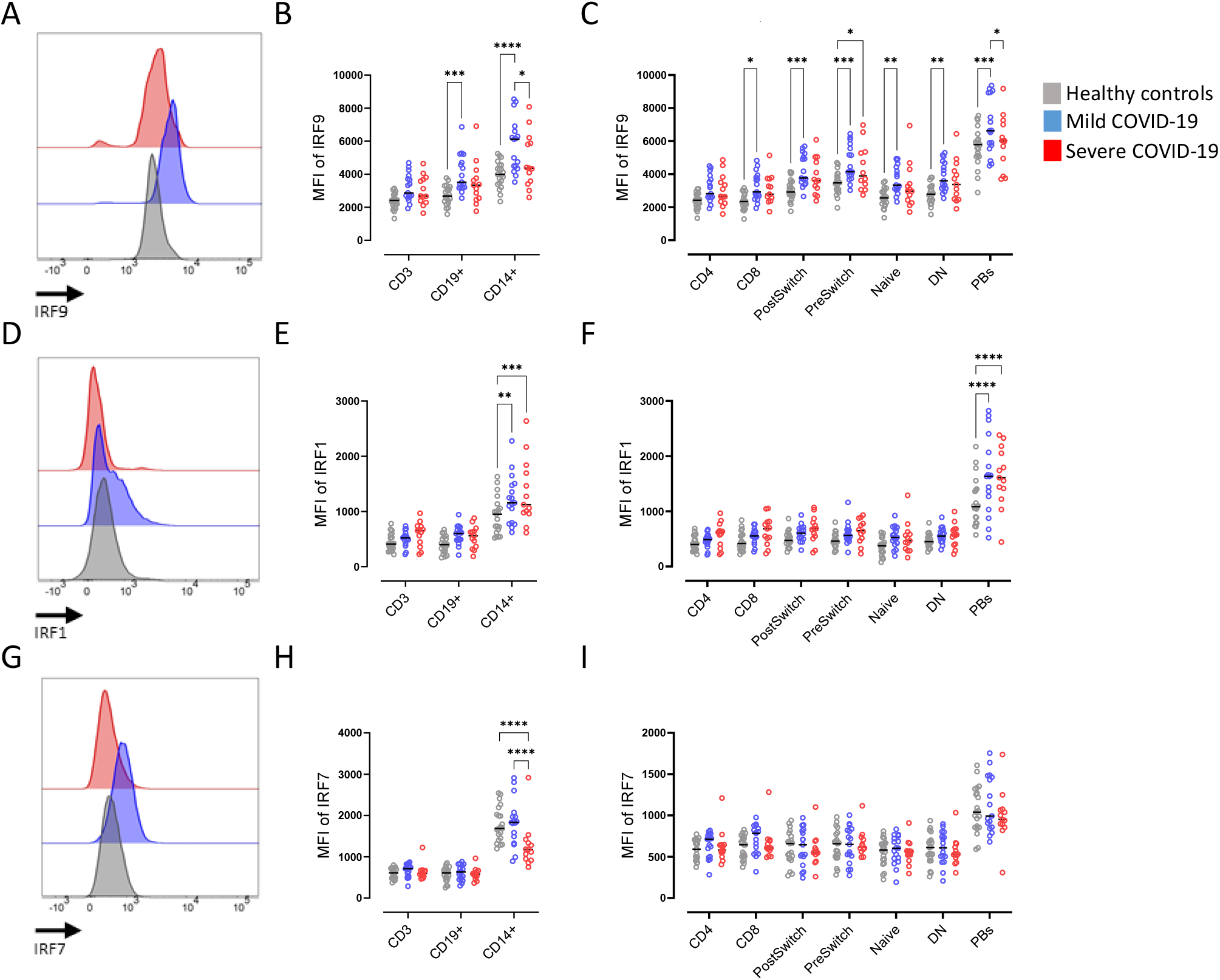
Enhanced intracellular IRF9 expression in severe COVID-19. (A) Representative histograms of baseline expression of IRF9 on B cells from healthy controls (grey), mild (blue) and severe (red) COVID-19 patients. (B) Median fluorescence intensity (MFI) of IRF9 in CD3+, CD19+ and CD14+ cells. (C) MFI of IRF9 in T and B cell subsets (as described in Fig1B). (D) Representative histograms of baseline expression of IRF1 on B cells healthy controls (grey), mild (blue) and severe (red) COVID-19 patients. (E) MFI of IRF1 in CD3+, CD19+ and CD14+ cells. (F) MFI of IRF1 in T and B cell subsets (as described in Figure 1B). (G) Representative histograms of baseline expression of IRF7 on B cells from healthy controls (grey), mild (blue) and severe (red) COVID-19 patients. (H) MFI of IRF7 in CD3+, CD19+ and CD14+ cells. (I) MFI of IRF7 in T and B cell subsets (as described in Figure 1B). Median and data from healthy controls (n=20), mild COVID-19 (n=17) and severe COVID-19 (n=13) patients. Two way ANOVA with Sidack post-test. *p<0.05, **p<0.01, ***p<0.001 and ****p<0.0001.

Expression of intracellular phosphorylated (pS477/pS479) IRF7 did not significantly differ between all groups studied and between the different T and B cell populations (Fig. 2 G-I). Only in monocytes, IRF7 was reduced in severely affected COVID-19 patients compared to mild cases and healthy controls.

### 3.4. Correlation of STAT1 with IFN signature

We wondered how these findings interrelate especially with regard to the IFN-signature, which was assessed by the expression of the surrogate marker Siglec-1 (CD169) on monocytes [20]. In our cohort we observed a significant reduction of Siglec-1 expression in severely affected COVID-19 patients (Figure 3A). Siglec-1 and STAT1 showed a significant correlation with in CD3+ T cells, CD19+ B cells and monocytes (Figure 3B), while pSTAT1 did not significantly correlate with Siglec-1 as a surrogate for IFN signature or e.g. age (Figure 3C). Also STAT1 and IRF9 both components of the ISGFR3 did not correlate (Figure 3C).

**Figure 3.**
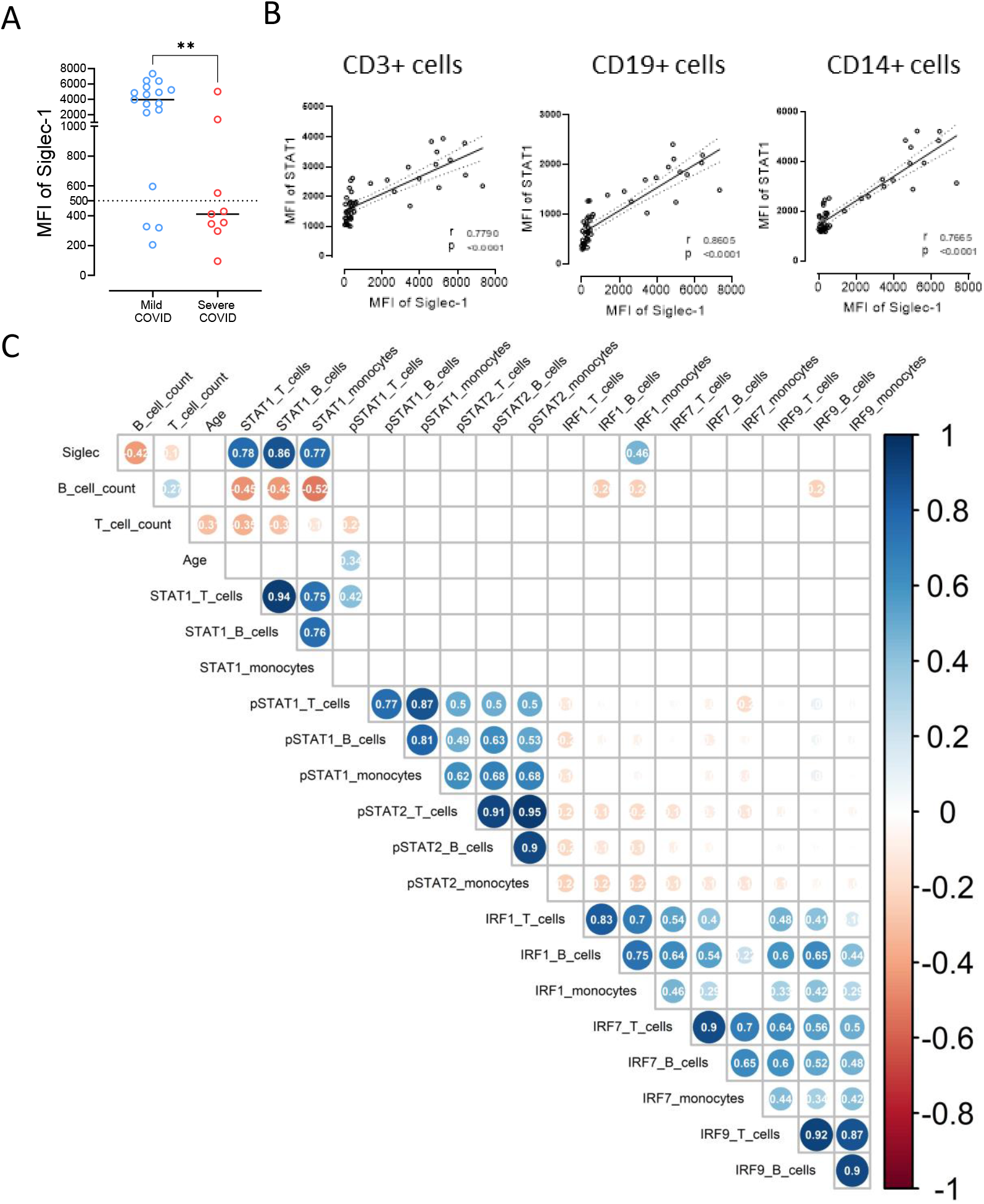
Significant correlation of STAT1 and Siglec-1 (CD169) (A) Siglec-1 (CD169) expression on CD14+ monocytes in mild and severe COVID-19 patients. (B) Correlation of Siglec-1 expression on the surface of CD14+ monocytes with intracellular STAT1 expression in CD3+ T cells, CD19+ B cells or CD14 + monocytes. Each point represents a donor. Mann-Whitney U test. *p<0.05, **p<0.01. (C) Spearman’s correlation matrix showing the correlation of all investigated parameters (STAT1, pSTAT1, pSTAT2, IRF1, IRF7 and IRF9) in relation to the analyzed cell populations (B cells, T cells and monocytes). Corresponding correlations are represented by red (negative) or blue (positive) circles. Size and intensity of color refer to the strength of correlation. Data from healthy controls (n=20), mild COVID-19 (n=17) and severe COVID-19 (n=13) patients. Only correlations with p ≤ 0.05 are indicated.

### 3.5. Reduced STAT1 response upon IFN stimulation in severe COVID-19

To further evaluate the functionality of type I and II IFN signaling in the context of our finding of reduced baseline whole protein expression of STAT1, we stimulated PBMCs of healthy controls, mildly and severely affected COVID-19 patients with low doses of either IFN-α or IFN-γ for 48h. As a control unstimulated cells (RPMI) of each group were used. Stimulation with IFN-α led to a transcriptional upregulation of STAT1 in CD3+ T cells and CD19+ B cells in healthy controls and in mild COVID-19 cases in CD19+ B cells (Fig 4A). In severe COVID-19 cases, a tendency of upregulation of STAT1 was observed (Figure 4A). The phosphorylation of STAT1 after low dose IFN-α or IFN-γ stimulation for 48 hours remained unchanged in healthy controls and mildly affected COVID-19 patients, while its was enhanced in severely affected COVID-19 patients already in unstimulated cells and without further increase after 48h incubation with IFN-α or IFN-γ (Figure 4B). Short term stimulation with higher doses of IFN-α was not able to increase pSTAT1 in severe COVID-19 cases. IFN-γ stimulation did not impact STAT1 phosphorylation (Figure 4C).

**Figure 4.**
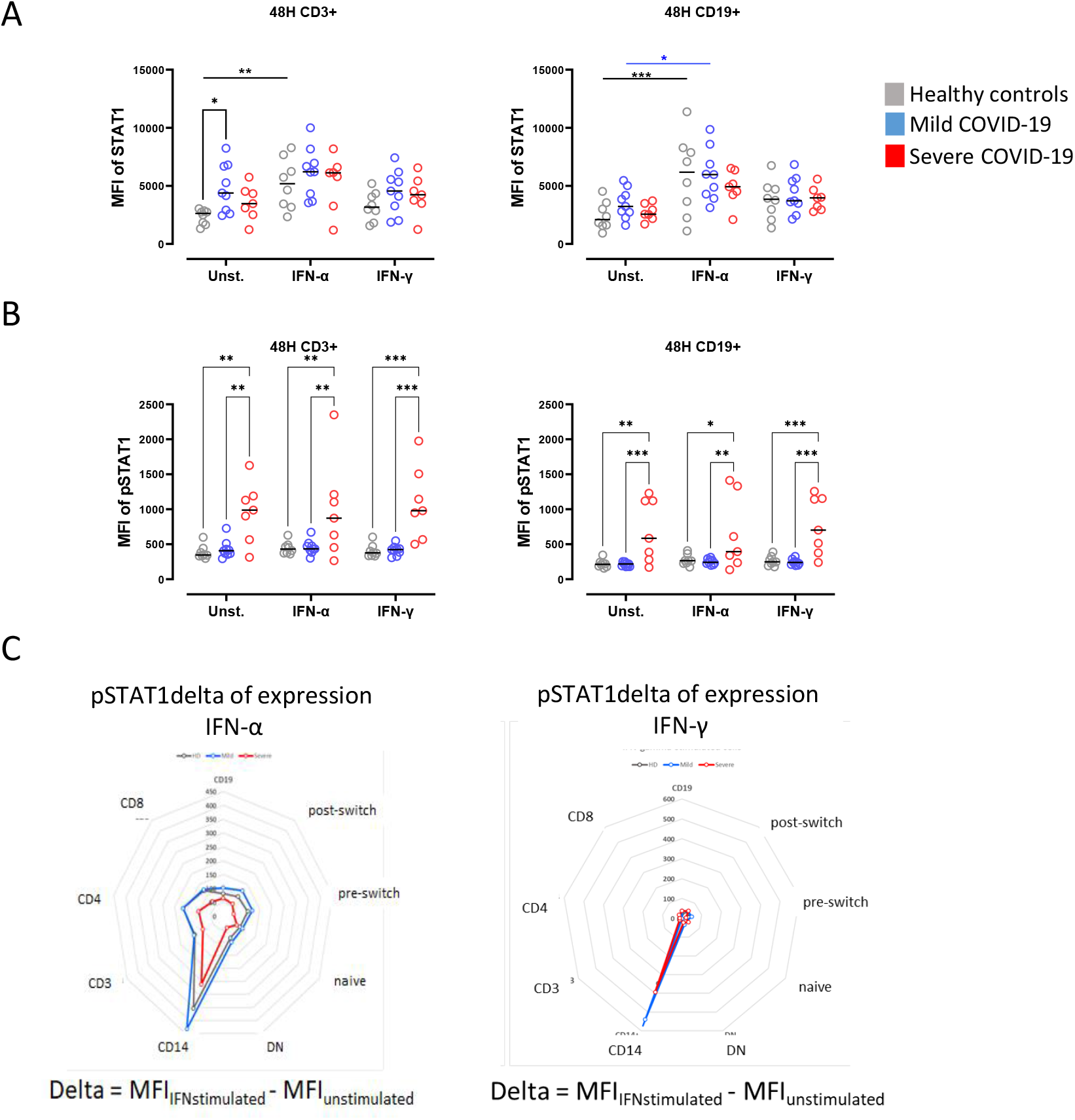
Attenuated pSTAT1 response upon IFN type I and II stimulation in severe COVID-19. (A) STAT1 and (B) pSTAT1 expression in CD3+ T cells and CD19+ B cells in culture of PBMCs from healthy controls (grey, n=8), mild (blue, n=9) or severe (red, n=7) COVID-19 patients. Cells were stimulated for 48 hours with either IFNα (5ng/ml) or IFNγ (5ng/ml) or only RPMI as a control. (C) PBMCs of the same donors as in (A) were stimulated with IFNα (100ng/ml) or IFNγ (100ng/ml) for 5 minutes. Untreated control values of pSTAT1 were subtracted to show the individual increase of STAT1 phosphorylation. Data is presented as radar diagrams. Two way ANOVA with Sidack post-test. *p<0.05, **p<0.01, ***p<0.001 and ****p<0.0001.

## 4. Discussion

COVID-19 is characterized by excessive production of multiple pro-inflammatory cytokines [21, 22] and patients with severe COVID-19 have a highly impaired IFN type I signature, in particular reduced IFN-α production and activity [6]. A common signaling route of cytokines, including IFNs is the JAK STAT signaling pathway [23]. JAKs and STATs provide a highly complex but orchestrated system of heterogeneous molecules with specific signaling through defined receptor complexes including recruitment of different STATs and resulting in specific downstream transcription [7].

In this study we describe an upregulation of STAT1 and IRF9 in mildly and severely affected COVID-19 patients, which correlated with the IFN-signature reflected by Siglec-1 surface expression. Both, Siglec-1 and STAT1 were lower among severely affected COVID-19 patients compared to mildly affected COVID-19 patients, especially in plasmablasts and monocytes. This is of particular interest since, these two cell types are considered the main players of pathogenesis in severe COVID-19 [24, 25]. The data suggest that certain viral factors may limit proper STAT1 and IRF9 function in severe COVID-19 patients in these cells and that the inhibition of translocation of STATs is more pronounced than the inhibition of phosphorylation [13, 14]. These results are in accordance with other authors, showing Siglec-1 expression correlate with viral load in mild COVID patients, but in severe COVID patients [26].

Increased levels of STAT1 in B cells were previously reported in SLE patients which correlate with Siglec-1 expression on CD14+ monocytes [18]. In a similar way, SLE patients have increased levels of the STAT1 protein in CD4+ T cells, alteration associated with perturbed homeostasis of regulatory T cells and disease severity [27]. Discordant to the whole protein levels, the phosphorylation of STAT1 is enhanced in severely affected COVID-19 patients suggesting a dysbalanced JAK STAT signaling that fails to induce transcription of the ISRE. An increased pSTAT1 is also present without stimulation in cultured PBMCS (here CD19+ B cells and CD3+ cells) and no further increase by IFN-α or IFN-γ stimulation is achieved. Further, this intervention could not demonstrate a transcriptional increase of STAT1 which would indicate reversibility of this condition.

Interestingly, phosphorylation of STAT1 in response to IFN stimulation can persist for hours, but newly synthesized unphosphorylated STAT1 induced by pSTAT1 can persist for several days [28]. This suggest high levels of unphosphorylated STAT1 in COVID patients were synthetized in response to persistent type I IFN signaling. It is worth to note that cells treated with lentivirus which exhibited increase in STAT1, did not completely phosphorylated in response to IFN stimulation [29]. Data shows that the antiviral role of STAT1 is not completely dependent of phosphorylation and suggests a role of alternative unphosphorylated STAT1 mediated pathways in COVID-19.

The reduced Siglec-1 expression on the surface of CD14+ monocytes of patients suffering from severe COVID-19 compared to patients with mild COVID-19 is consistent with published data [30-32]. As a result of this finding and efficient analysis of Siglec-1 (CD169) on CD14+ cells, we started to routinely measure Siglec-1 in all patients with COVID-19 admitted to ICUs to distinguish between patients with high and low interferon signature to test the hypothesis of an association of IFN signature and outcome of the patients.

Absence of correlation between STAT1 and pSTAT1 with IRF9, suggest that an alternative IRF9 independent signaling [12] could have a role in COVID-19. Previously, it was reported for IRF9 knock-out mice that type I IFN mediates a potent inflammatory response associated with a more severe neurological disease [33]. Varicella zoster virus prevents type I IFN response reducing IRF9 and inhibition of STAT2 phosphorylation in infected cells [34]. IRF9 also prevents exhaustion of CD8+ T cells in Lymphocytic choriomeningitis infection [35]. Interestingly, SARS-CoV-2 spike transfected cells secrete miR-148a and −590 via exosomes which induce degradation of IRF9 in human microglia [36].

Currently, optimal treatment for patients with COVID-19 is still uncertain and both interventions, blocking of IFN signaling by JAK STAT inhibition [37, 38] or the use of different types of IFN as substitutions have been suggested as a treatment for COVID-19 and showed efficacy in small clinical trials [39, 40]. Thus, the key question raises which intervention is appropriate for which patient? In patients with high IFN signaling that are likely in an early stage of the disease, inhibition strategies appear to be attractive especially during the phase of cytokine storm. On the other hand, patients with inability to increase antiviral response are likely in severe stage of disease, IFN substitution may hold more promise than further inhibition.

The current data indicate severe impairment of STAT1 and IRF9 in severely infected COVID-19 patients consistent with inappropriate type IFN upregulation as a potential mechanism for enhanced disease severity. It appears of utmost importance to understand the underlying mechanisms in more detail as patients with diminished type IFN response may benefit from targeted therapies. In this regard, JAK STAT inhibition may hold promise for patients with higher STAT1 expression (mild cases), while patients with low Siglec-1 and unable inability to increase antiviral response may benefit from IFN substitution.

## Data Availability

Data are available on reasonable request.

## 6. Ethics Statement

This study was carried out in accordance with the recommendations of the ethics’ committees at the Charité University Hospital Berlin and University of Ulm with written informed consent from all subjects. All subjects gave written informed consent in accordance with the Declaration of Helsinki.

## 7. Author Contributions

ES, AA, JR, DK, DZ and LS enrolled patients and collected samples.

HR, ES, FSZ, LS and AA analyzed the data.

SK, HS provided patients within the clinical trial.

KUE, TD, AL, ES and HS supervised the work and acquired funding

All authors developed, read, and approved the current manuscript.

---

**Fig. S1.**
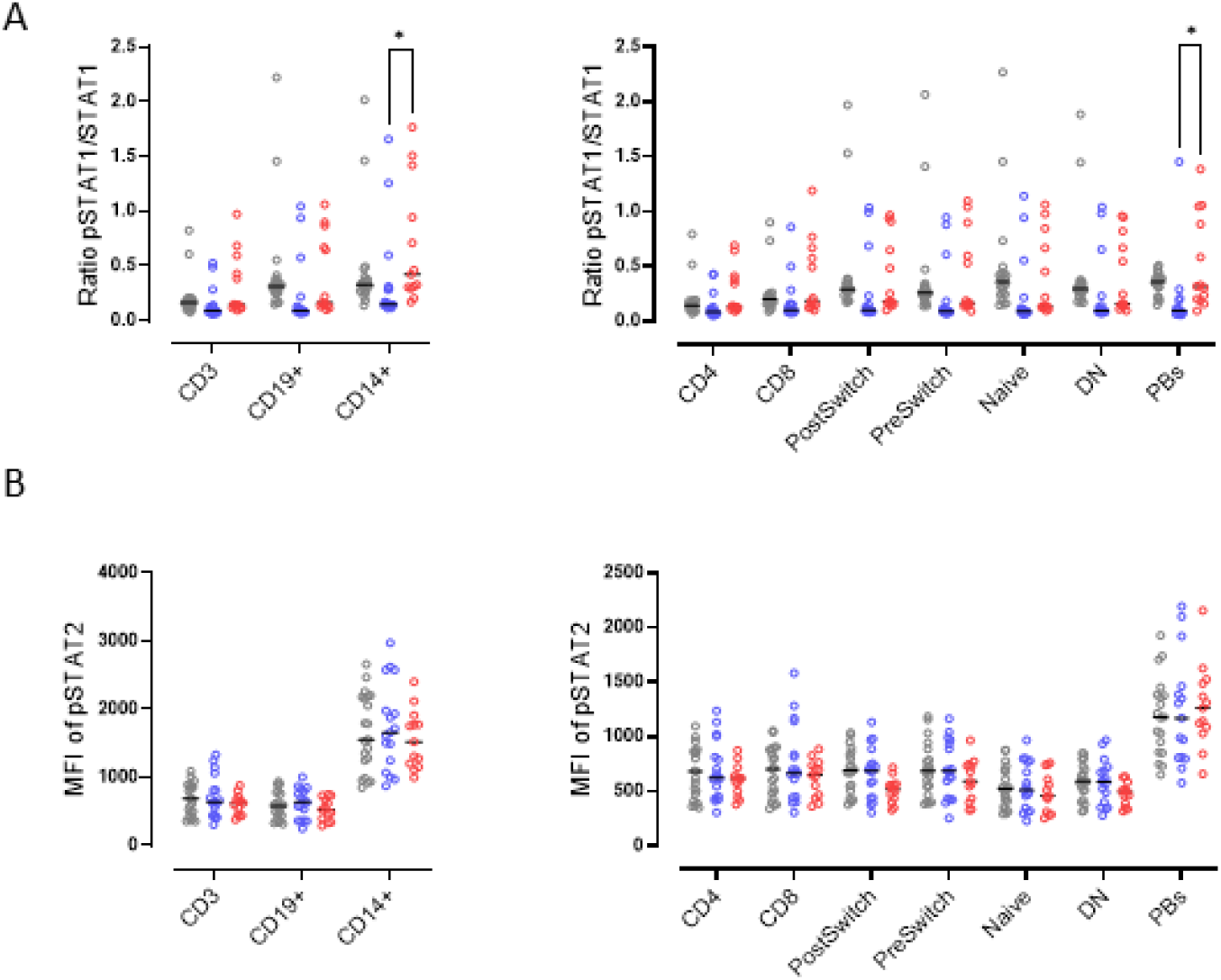
Intracellular pSTAT2 levels and pSTAT1/STAT1 ratio of HD and COVID-19 patients. **(A)** Ratio of pSTAT1/STAT1 in CD3+, CD19+ and CD14+ cells. Ratio of pSTAT1/STAT1 in T and B cell subsets (as described in Fig1B). **(B)** Median fluorescence intensity (MFI) of pSTAT2 in CD3+, CD19+ and CD14+ cells. MFI of pSTAT2 in T and B cell subsets (as described in Figure 1B). Median and data from HD (n=21), mild COVID-19 (n=17) and severe COVID-19 (n=13) patients. Two way ANOVA with Sidack post-test. *p<0.05, **p<0.01, ***p<0.001 and ****p<0.0001.

